# OVARIAN GRANULOSA CELLS FROM WOMEN WITH PCOS EXPRESS LOW LEVELS OF SARS-COV-2 RECEPTORS AND CO-FACTORS

**DOI:** 10.1101/2021.06.15.21259003

**Authors:** Aalaap Naigaonkar, Krutika Patil, Shaini Joseph, Indira Hinduja, Srabani Mukherjee

## Abstract

**Purpose:** Severe acute respiratory syndrome coronavirus-2 (SARS-CoV-2) infection is global pandemic with more than 3 million deaths so far. Female reproductive tract organs express coronavirus-associated receptors and factors (SCARFs); suggesting they may be susceptible to SARS-CoV-2 infection however the susceptibility of ovary/follicle/oocyte to the same is still elusive. Co-morbidities like obesity, type-2 diabetes mellitus, cardiovascular disease etc. increase the risk of SARS-CoV-2 infection. These features are common in women with polycystic ovary syndrome (PCOS), warranting further scope to study SCARFs expression in ovary of these women.

**Materials and methods:** SCARFs expression in ovary and ovarian tissues of women with PCOS and healthy women was explored by analyzing publically available microarray datasets. Transcript expression of SCARFs were investigated in mural and cumulus granulosa cells (MGCs and CGCs) from control and PCOS women undergoing in vitro fertilization (IVF).

**Results:** Microarray data revealed that ovary expresses all genes necessary for SARS-CoV-2 infection. PCOS women mostly showed down-regulated/unchanged levels of SCARFs. MGCs and CGCs from PCOS women showed lower expression of receptors *ACE2, BSG* and *DPP4* and protease *CTSB* than in controls. MGCs showed lower expression of protease *CTSL* in PCOS than in controls. Expression of *TMPRSS2* was not detected in both cell types.

**Conclusions:** Human ovarian follicle may be susceptible to SARS-CoV-2 infection. Lower expression of SCARFs in PCOS indicate that the risk of SARS-CoV-2 infection to the ovary may be lesser in these women than controls. This knowledge may help in safe practices at IVF settings in the current pandemic.

## Introduction

Severe Acute Respiratory Syndrome Coronavirus-2 (SARS-CoV-2) causes the infection Coronavirus disease 2019 (COVID-19) which is currently a public health emergency worldwide. The disease in symptomic patients shows a broad spectrum of clinical manifestations ranging from fever, cough, tiredness, joint/muscle pain, shortness of breath, headache, hemoptysis, diarrhea, dyspnea etc. [1]. Since the first case in December 2019, SARS-CoV-2 has infected more than 170 million individuals from 215 countries and territories as of June 16^th^ 2021, leading to more than 3.8 million deaths with an average mortality rate of around 4% (WHO, 2021).

The coronavirus is a large group of enveloped, single-strand positive-sense RNA viruses classified as beta coronavirus. Genetic sequence of SARS-CoV-2 shares at least 70% homology with SARS-CoV and around 50% with the MERS-CoV [2]. The recent outbreak of COVID-19 is much more contagious than SARS-CoV. Despite the approval of few vaccines for SARS-Cov-2, it would still be challenging to control the pandemic considering various factors like the speed at which vaccines are developed, administered; also its administration to high risk and vulnerable groups like pregnant women and individuals below 18 years of age, complex social policies and the acceptance of vaccine by the population etc. Besides, the emerging mutations in SARS-CoV-2 genome may enable it with increased infectivity and virulence, immune evasion and impedance against diagnostics and therapy hence raising the concerns of scientific and medical community to a greater extent [3].

Molecular features of these coronavirus family members have been decoded well. The information on these molecular determinants help us to understand the mechanism of infection which involves both viral and host cellular machinery. There are around 28 SARS-CoV-2 and coronavirus-associated receptors and factors (SCARFs) that can be attributed with different roles in SARS-CoV-2 infection and pathogenesis, out of which we are hereby discussing that are in the interest of this study [4]. The viral RNA is encapsulated in envelope, that consists of three proteins; viz. envelope protein, membrane protein and spike protein (S protein). The transmembrane glycoprotein, S protein which is involved in binding and infect host cell is responsible for variation in corona virus and host tropism [5]. S protein binds to the host cells by the canonical receptor, angiotensin converting enzyme II (ACE2). The virus may also bind to Basigin (BSG) (non-canonical receptor) through S protein [4]. Dipeptidyl-peptidase 4 (DPP4) and C-Type Lectin Domain Family 4 Member M (CLEC4M) are other molecules proposed to be used by virus for entry into the host cell [6, 7]. The receptor attachment is followed by priming of S protein by host proteases to facilitate viral entry. Transmembrane protease, serine 2 (TMPRSS2) mediates entry of the virus into the host cells by cleaving the S protein followed by membrane fusion. In the absence of TMPRSS2, the virus can use cathepsin B/L (CTSB and CTSL), FURIN etc. as alternate proteases for the priming of the S protein [4].

Although pulmonary damage has been the most prevalent cause of death due to COVID-19 there is growing evidence for the emerging multi-organ infectious nature of SARS-CoV-2 [4] which warrants studying the susceptibility of other organs to the infection. Apart from lungs, COVID-19 has been reported to affect human heart, liver, kidney and nervous system as well [8]. Multi-organ map for expression of SCARFs has been dissected [9]. Theoretically, SARS-CoV-2 may infect any cell or tissue type that co-expresses receptor and proteases, i.e. ACE2 and TMPRSS2 [10]. There are reports confirming the presence of ACE2 expression at ovary, uterus, vagina and placenta [9, 11–14]. In addition to this, expression of other receptors and proteases (BSG, CTSL, FURIN) has been demonstrated in male and female reproductive tissue by several studies by using single cell RNA sequencing approach and by analyzing publically available microarray datasets [4, 15]. On the other hand, TMPRSS2 expression in ovary is inconsistent but alternative proteases still remain in contention to facilitate the SARS-CoV-2 infection at ovary. Zhou et al performed in-situ protein proofed single cell RNA profiling of several human tissues which indicated that ovary can be a likely target for SARS-CoV-2 infection [16]. Initially it was thought that placenta do not express receptors of SARS-CoV-2 but several studies have confirmed the SCARFs expression at feto-maternal interface [17–19].

The susceptibility of the human oocyte/follicle to SARS-CoV-2 infection however still remains elusive. If this is possible then it may in turn affect the oocyte quality, ovulation, fertilization, implantataion and may even pose the risk to the growing embryo outcome [20]. ESHRE (European Society of Human Reproduction and Embryology) in March 2020 advocated for avoidance of pregnancy and also not to commence infertility treatment to avoid further stress on strained healthcare system in pandemic [21]. Thereafter, to gradually start with ART (assisted reproductive technique) clinics, ESHRE and ASRM (American Society for Reproductive Medicine) laid down guidelines for careful practices of oocyte retrieval, embryo transfer, cryopreservation and for general safety in ART clinics as there is a possibility of contamination through culture media, equipment or working personnel infected with SARS-Co-2 [22]. It is still not clear whether the gametes and embryos are susceptible to SARS-Co-2 infection in vitro. It is thus crucial to know about expression of receptors and molecules that can effectuate SARS-CoV-2 infection in these cells.

Oocytes of non-human primate have also showed ACE2 expression along with the expression of co-receptor BSG and alternative protease CTSL [10]. Studies showed expression of both *ACE2* and *BSG* receptors in human oocytes and preimplantation embryos (blastocyst) [20, 23]. In the follicle, oocyte is surrounded by companion somatic cells i.e. cumulus granulosa cells (CGCs) whereas the mural granulosa cells (MGCs) line the follicular wall around the antrum. While MGCs are involved in more of steroidal functions and providing mechanical support to oocyte, the CGCs are in immediate contact with oocyte and act as metabolic drivers of the oocyte [24]. As MGCs and CGCs are in close vicinity of oocyte, if they are susceptible to infection by SARS-CoV-2 there are high chances that the oocyte may also get infected. Hence it is important to know about SCARFs expression on these cells which would indicate the susceptibility of oocyte and in turn the embryo towards SARS-CoV-2 infection. However, SCARFs expression in granulosa cells has not been studied much apart from a few reports [10, 25] hence there is a need to investigate SCARFs expression in ovarian cells. Therefore, we investigated the expression of receptor and proteases (which facilitate the entry of SARS-CoV-2 inside the cells) in ovarian granulosa cells, from women undergoing in vitro fertilization (IVF).

Further, the risk of SARS-CoV-2 infection varies in people with different health issues. It has been reported that individuals with pre-existing co-morbidities like obesity, type-2 diabetes mellitus (T2D), cardiovascular disease (CVD) are more prone to SARS-CoV-2 infection [26]. These clinical manifestations are presented as co-morbidities in polycystic ovary syndrome (PCOS) which is the most common endocrine disorder in women of reproductive age with the incidence rate of 6-15% [27]. This overlap between cardio-metabolic profiles of women with PCOS may pose them at high risk of COVID-19 and may even worsen clinical outcomes of COVID-19. Additionally, hyperandrogenism in these women may lead to elevated ACE expression which augments viral entry [27]. Recent population based study by Subramanian reports increased COVID-19 risk in women with PCOS due to presence of co-morbidities [28]. Moreover, there are evidences showing that the receptors for SARS-CoV-2 (ACE2, TMPRSS2) can be regulated by androgens, which are elevated in PCOS [29]. BSG too may be regulated by estrogen and progestertone [30]. However, the possible influence or chances of infection to follicles/ oocyte which may affect the fertility/ pregnancy and its outcome due to COVID-19 in PCOS women is still not clear.

Hence, we first explored the expression of SCARFs in ovarian tissue of healthy women and women with PCOS by analyzing publically available datasets of microarray studies. Also, we studied transcript expression of SCARFs in MGCs and CGCs of women with PCOS and control women which has not been studied as much till date.

## Materials and methods

### 1. Microarray data analysis

The Gene Expression Omnibus (GEO, http://www. http://ncbi.nlm.nih.gov/geo/) database was searched for gene expression datasets on PCOS compared to controls (Table 1). Seven PCOS datasets which compared gene expression patterns of ovarian cells/tissues in control women and women with PCOS were selected and downloaded from the GEO database [31]. Each of the datasets was processed independently to identify differentially expressed genes. The microarray data analysis was carried using R 4.0.2. The raw CEL files for the Affymetrix arrays were downloaded and normalized using the robust multiarray average method (RMA) to allow for background correction, normalization and summarization. In case of the Agilent arrays, the pre-processed quantile normalized series matrix file were used. After preprocessing, the unmapped probes were removed. Duplicate gene entries were collapsed using the collapse Row function in R. The limma package in R was used to identify differentially expressed genes [32]. Multiple testing correction was performed using the Benjamini and Hochberg method. Genes with p-value <0.05 and fold change >1.5 were considered to be differentially expressed.

**Table 1:** Gene expression datasets included in the study. List of microarray datasets downloaded from GEO. The studies were carried out in women with PCOS and controls were used for the analysis using R package.

| **Dataset** | **Sample used for analysis** | **Platform** | **Array** |
| --- | --- | --- | --- |
| GSE114419 | Controls (3)<br>PCOS (3) | GPL17586 | Affymetrix Human Transcriptome Array 2.0 |
| GSE137684 | Controls (4)<br>Normo-androgenic PCOS (3) | GPL17077 | Agilent-039494 SurePrint G3 Human GE v2 8x60K Microarray 039381 |
| GSE106724 | Controls (4)<br>Normo-androgenic PCOS (4) | GPL21096 | Agilent-062918 Human lncRNA array V4.0 |
| GSE102293 | Controls (4)<br>PCOS (2) | GPL570 | Affymetrix Human Genome U133 Plus 2.0 Array |
| GSE34526 | Controls (3)<br>PCOS (7) | GPL570 | Affymetrix Human Genome U133 Plus 2.0 Array |
| GSE5850 | Controls (6)<br>PCOS (6) | GPL570 | Affymetrix Human Genome U133 Plus 2.0 Array |
| GSE1615 | Controls (4)<br>PCOS (5) | GPL96 | Affymetrix Human Genome U133A Array |

### 2. Study subjects and sample collection

This study was carried out at ICMR-National Institute for Research in Reproductive Health (ICMR-NIRRH), India after ethical approval. We recruited women with PCOS (n=14) as per the Rotterdam consensus criteria [33]. Regularly menstruating women having no reproductive complications and undergoing IVF due to male factor infertility or oocyte donors were recruited as controls (n=15). All participants were undergoing controlled ovarian hyperstimulation using a GnRH agonist protocol for IVF at P. D. Hinduja National Hospital and Medical Research Centre, Mumbai. They were enrolled in the study after obtaining written consent. On the day of ovum pick up (d-OPU), blood was collected from all participants for carrying out biochemical and hormonal assays. On the same day macroscopically clear follicular fluid was collected, processed as described previously [34]. Serum and follicular fluid collected on d-OPU were assayed for estradiol (E_2_), progesterone (P_4_), total testosterone (TT) and SHBG by electro-chemiluminescence technology using Roche e411 automated analyser (Roche, Basel, Switzerland). Baseline levels for LH, FSH, prolactin and TSH estimated between days 3–7 of menstrual cycle were obtained from clinical records. TT and SHBG values were used to calculate androgen excess indices (http://www.issam.ch/freetesto.htm).

### 3. Isolation of granulosa cells

CGCs were separated manually from the cumulus oocyte complex suspended in aspirated follicular fluid. From the IVF center, CGCs were transported in ovum buffer to the lab for further processing. CGCs were then washed with PBS and cell lysis buffer was added to extract RNA. MGCs were collected and separated from red blood cells by centrifugation through a ficoll gradient (HiMedia, India) at 600g for 20 mins. MGCs were carefully removed from the middle layer of ficoll gradient and washed with phosphate-buffered saline. The enriched CGCs and MGCs were used for quantitative gene expression analysis.

### 4. Real-Time PCR

RNA was extracted from cells using Qiagen miRNA easy kit (Qiagen, Hilden, Germany) and quantified by Nanodrop Synergy HT (Biotek, Germany). The cDNA was synthesized by high-capacity cDNA reverse transcription kit (Applied Biosystems, CA, USA). The expression of genes was investigated using the Takyon SYBR mastermix (Eurogentec, Europe) and appropriate primers (Supplemental Table 1) using cDNA samples. These included receptors *ACE2, BSG, CLEC4M, DPP4* and proteases *TMPRSS2, CTSB, CTSL* and *FURIN*. The mRNA abundance was normalized to the expression of housekeeping gene 18S rRNA, and the gene expression levels were represented as fold change values by the Δ threshold cycle (Ct) 2^-ΔΔCt^ method.

## Results

### 1. Clinical characteristics of the study participants

There was no significant difference between age and BMI between control and PCOS group (Table 2). On the d-OPU, P4 levels in follicular fluid and SHBG levels both in follicular fluid and serum of women with PCOS were significantly lower than controls. Basal LH and LH:FSH ratio in serum and total, bioavailable and free testosterone as well as free androgen index were significantly higher in follicular fluid and serum collected on d-OPU in PCOS group as compared to control group.

**Table 2:** Demographic and clinical characteristics of the study participants undergoing controlled ovarian hyperstimulation during IVF. Data are represented as the median (inter-quartile range) for demographic, hormonal, and biochemical profiles compared between control and women with PCOS. Parameters marked with “**$”** were measured in serum and follicular fluids obtained on the day of ovum pick up. Statistical comparison was performed using the Mann-Whitney U test. *P* values < 0.05 are considered significant for all statistical tests. \**P* < 0.05, \*\**P* < 0.01. IVF, in vitro fertilization; BMI, body mass index; LH, luteinizing hormone; FSH, follicle stimulating hormone; TSH, thyroid stimulating hormone; FF, follicular fluid; E_2_, estradiol; P_4,_ progesterone; TT, total testosterone; SHBG, sex hormone binding globulin; Free T, free testosterone; Bio-T, bioavailable testosterone; FAI, free androgen index.

|  | Control (n=15), Median (IQR) | PCOS (n=14), Median (IQR) | *P* value |
| --- | --- | --- | --- |
| Age, years | 28 (22-31) | 28.50 (25.75-31) | 0.430 |
| BMI (kg/m<sup>2</sup>) | 20.50 (18.40-25.40) | 24.50 (21.58-28.26) | 0.077 |
| Basal LH levels (μU/mL) | 3.6 (2.8-6.13) | 7.57 (4.9-8.995) | **0.0007\*\*** |
| Basal FSH levels (μU/mL) | 5 (4.48-7.09) | 5.65 (4.11-6.14) | 0.810 |
| LH:FSH | 0.656 (0.54-0.9) | 1.37 (1.04-1.65) | **0.0006\*\*** |
| Prolactin (ng/mL) | 14.2 (6.37-16.98) | 12.3 (10.18-14.56) | 0.743 |
| TSH (mIU/mL) | 1.87 (1.6-3.08) | 2.08 (1.29-3.20) | 0.777 |
| E2 (ng/mL) before hCG administration | 1.68 (1.5-2.24) | 1.79 (1.22-2.4) | 0.948 |
| E2 (ng/mL) on hCG administration day | 2.49 (2.1-3.8) | 2.54 (2.25-4.19) | 0.694 |
| <sup>\$</sup>E<sub>2</sub> (ng/mL) Serum | 2.14 (2-2.8) | 0.997 (0.078-2.61) | 0.067 |
| <sup>\$</sup>E<sub>2</sub> (ng/mL) FF | 840 (435-1042) | 912.6 (608.9-1260) | 0.420 |
| <sup>\$</sup>P<sub>4</sub> (ng/mL) Serum | 2.6 (1-5) | 0.73 (0.188-2.98) | 0.073 |
| <sup>\$</sup>P<sub>4</sub> (μg/mL) FF | 23.02 (15-30) | 15.48 (7.03-21.06) | **0.036\*** |
| <sup>\$</sup>TT (ng/dL) Serum | 111 (90-128) | 161.5 (123.7-192.5) | **0.019\*** |
| <sup>\$</sup>TT (ng/dL) FF | 372.6 (260-600) | 617.5 (400.3-787.4) | **0.025\*** |
| <sup>\$</sup>SHBG (nmol/L) Serum | 125 (80-234) | 89.85 (47.25-108.4) | **0.045\*** |
| <sup>\$</sup>SHBG (nmol/L) FF | 153.4 (133.2-175) | 95.55 (78.25-149.9) | **0.034\*** |
| <sup>\$</sup>Free T (pmol/L) Serum | 28.10 (19.5-34) | 52.87 (36.37-61.75) | **0.001\*\*** |
| <sup>\$</sup>Free T (pmol/L) FF | 87.09 (58-127) | 183 (142.8-224.5) | **0.0005\*\*** |
| <sup>\$</sup>Bio T (nmol/L) Serum | 0.66 (0.46-0.79) | 1.24 (0.85-1.45) | **0.001\*\*** |
| <sup>\$</sup>Bio-T (nmol/L) FF | 2.09 (1.35-2.98) | 4.3 (3.34-4.93) | **0.0005\*\*** |
| <sup>\$</sup>FAI Serum | 3.28 (2.26-4.25) | 6.24 (4.27-8.88) | **0.001\*\*** |
| <sup>\$</sup>FAI FF | 7.66 (6.44-13.15) | 19.50 (16.29-24.41) | **0.0004\*\*** |

### 2. Microarray data analysis

We carried out microarray data analysis using gene expression datasets on studies involving PCOS and control women (Fig1). Interestingly, all SCARFs which we have focused in the present study are expressed in the ovary of healthy women. Between controls and PCOS, majorly of the COVID related host genes did not show any differential expression except few. Analysis showed *ACE2* was down-regulated in granulosa cells of women with PCOS compared to controls [35]. Another receptor, *BSG* showed lower expression in theca cells but in granulosa cells its expression was increased in PCOS women than in controls [36, 37]. DPP4 was found to be decreased in granulosa cells of women with PCOS [37]. Further, *FURIN* was observed to be down-regulated in oocyte and up-regulated in granulosa cells of women with PCOS [35, 38]. *CTSB* expression was observed to be up-regulated in both oocyte and granulosa cells of women with PCOS [37, 38]. *CTSL* expression was not consistent in granulosa cells of PCOS across different microarray studies (GSE106724), [37, 39]. The expression of *CLEC4M* and *TMPRSS2* were comparable across all analyzed studies. However, the microarray data for these genes are not validated by gene expression analysis.

**Fig1:**
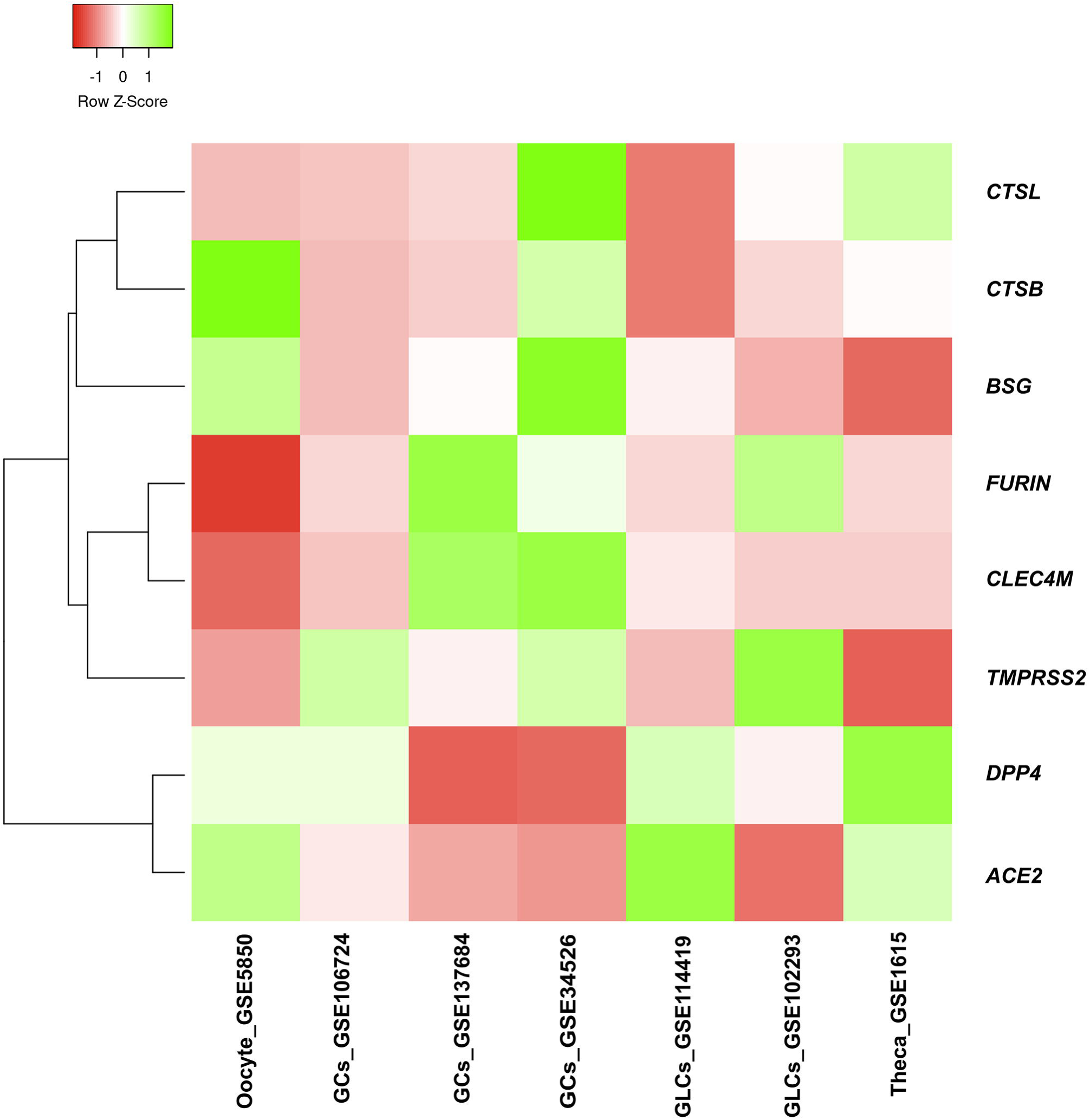
Heat map showing the expression of genes for SARS-CoV-2 receptors and proteases in ovary and ovarian tissues from women with PCOS compared to controls. Differential expression of genes for SARS-CoV-2 receptors and spike protein processing enzymes in ovary from microarray expression gene datasets obtained from GEO. *ACE2*, angiotensin converting enzyme II; *BSG*, basigin; *DPP4*, dipeptidyl-peptidase 4; *CLEC4M*, C-type lectin domain family 4 member M; *TMPRSS2*, transmembrane protease, serine 2; *CTSB*, cathepsin B; *CTSL*, cathepsin L; GCs, granulosa cells; GLCs, granulosa-Lutein cells.

### 3. Transcript levels of SCARFs in MGCs and CGCs

We measured the transcript levels of several SARS-CoV-2 receptors (*ACE2, BSG, CLEC4M, DPP4*) and proteases (*TMPRSS2, CTSL, CTSB, FURIN)* and found all of them are expressed in ovarian granulosa cells (both MGC and CGCs) except *TMPRSS2* (Fig2). In women with PCOS, most gene expression pattern of SCARFs in MGCs and CGCs accorded with each other. The expression of receptors *ACE2, BSG* and co-receptor *DPP4* was significantly lower but *CLEC4M* transcript level was comparable in women with PCOS and controls in both MGCs and CGCs. In case of proteases transcript levels of both *CTSB* and *CTSL* were significantly lower in MGC whereas only *CTSL* transcript level was decreased in CGCs of women with PCOS compared to controls. Expression of *FURIN* was comparable between control and women with PCOS in both MGC and CGC.

**Fig2:**
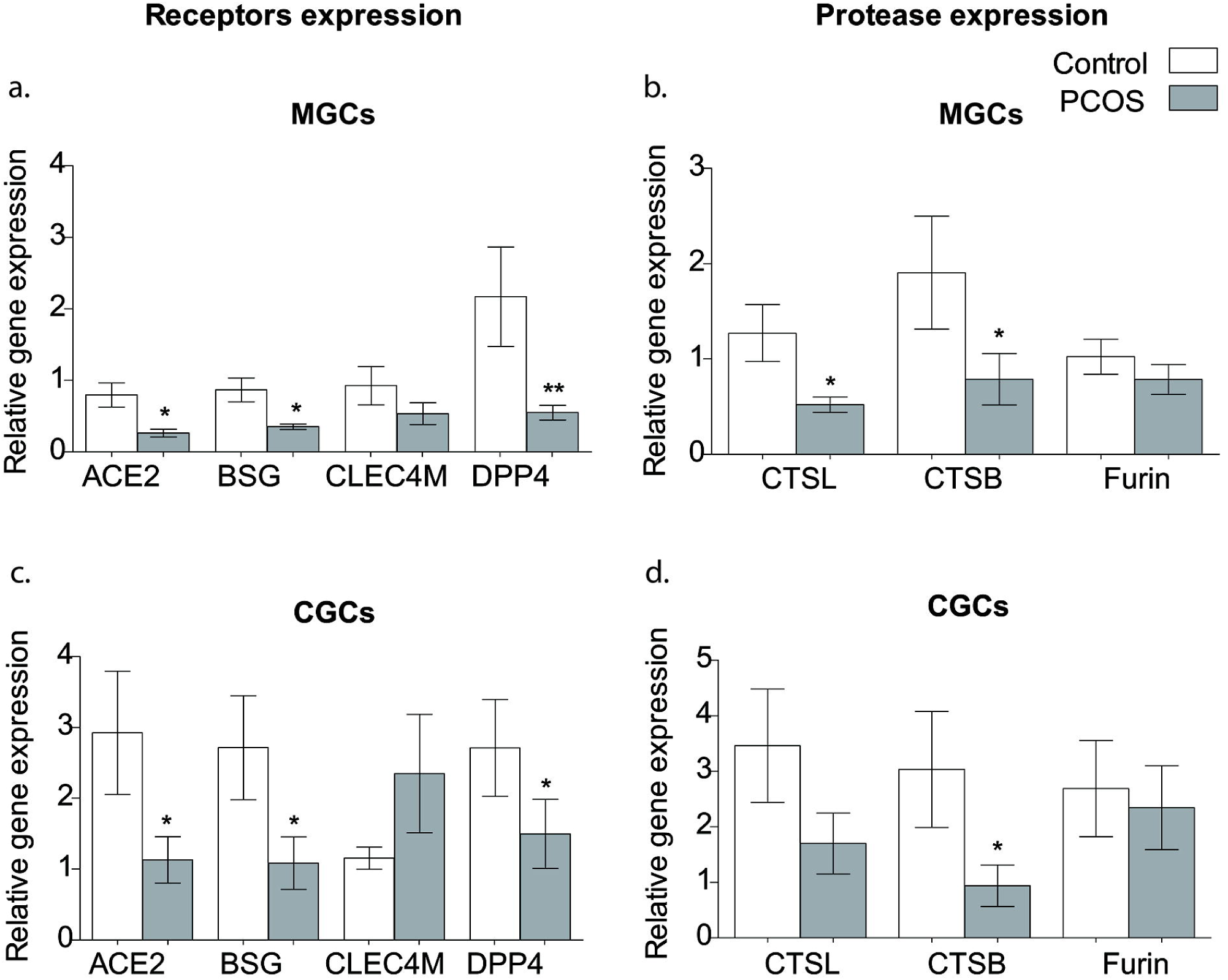
Relative expression of SCARFs in granulosa cells. Relative genes expression levels of SARS-CoV-2 receptors (*ACE2, BSG, CLEC4M, DPP4*) and spike protein processing enzymes (*CTSB, CTSL and FURIN*) in MGCs (a and b) and CGCs (c and d) compared between PCOS and control groups. Fold change was evaluated using the 2^-ΔΔCt^ method. Expression was normalized to the 18S rRNA gene as an endogenous control and granulosa cells calibrator sample. Bar graphs represent “mean ± SEM” and *P < 0.05 considered significant. Data are analyzed using the Mann-Whitney U test. MGCs, mural granulosa cells; CGCs, cumulus granulosa cells; *ACE2*, angiotensin converting enzyme II; *BSG*, Basigin; *DPP4*, Dipeptidyl-peptidase 4; *CLEC4M*, C-Type Lectin Domain Family 4 Member M; *TMPRSS2*, Transmembrane protease, serine 2; *CTSB*, cathepsin B; *CTSL*, cathepsin L.

## Discussion

COVID-19, one of the most threatening zoonotic outbreaks seen so far, affects respiratory tract majorly. However recent evidences have indicated that there is a possibility of infection to other organs [4]. Considering this, the organs of reproductive system have got attention as infection to them may in turn negatively influence production and maturation of gametes, fertilization and pregnancy [20]. Moreover, individuals with pre-existing co-morbidities like T2DM, CVD are more susceptible to SARS-CoV-2 infection and women with PCOS are at risk of development of diabetes, hypertension and CVD [26]. Infertility is common in women with PCOS so they often seek IVF for infertility treatment [40]. Infection to the ovary and granulosa cells may hamper fertilization and pregnancy outcomes during IVF procedures [41]. Taking all this into account, we have analyzed microarray datasets from the studies carried out in the ovary and ovarian tissue of women with PCOS and compared it to controls along with the validation of these genes in granulosa cells.

The analysis of microarray data revealed that the healthy ovary expresses all genes necessary for the SARS-CoV-2 infection indicates that the ovary might be susceptible to infection by SARS-CoV-2. The comparative analysis between controls and PCOS showed lower or unchanged receptor expression and either unchanged or higher proteases expression across all analyzed studies and the results are not consistent. However, in case of PCOS, all the receptors were down-regulated in ovary, hinting towards the reduced risk of ovarian infection in them.

Apart from the microarray studies, reports on the expression of SCARFs in MGCs and CGCs are rare [10, 25]. Therefore, we investigate the expression of SCARFs in granulosa cells of PCOS and controls and the susceptibility of the ovary/ovarian follicle to SARS-CoV-2 infection. The *ACE2* expression was downregulated in both CGCs and MGCs from women with PCOS in our study. This observation of lower *ACE2* corroborates with our microarray gene expression analysis of granulosa cells, plasma levels reported in PCOS women and studies on oocytes from rat model of PCOS [35, 42, 43]. Localization and expression of BSG receptor has been a point of interest considering that BSG is known to be regulated by estrogen and progesterone and is important for fertilization and implantation [30]. A study involving women with and without endometriosis reported expression of BSG at granulosa cells (follicles of all stages), ovarian surface epithelium, and corpora lutea [11]. Also, Essahib et al have confirmed the presence of BSG on oocyte membrane in women undergoing IVF [23]. *BSG* was found to be down-regulated in theca cells and up-regulated in granulosa cells of PCOS women compared to controls according to microarray analysis [36, 37]. On the contrary, we found significantly lower expression of BSG in MGCs and CGCs of women with PCOS. Our data on BSG expression in both types of granulosa cells further indicate that oocyte can be infected by SARS-CoV-2 however women with PCOS may be at lower risk of infection.

DPP4, co-receptor may also play a major role in entry and virulence of SARS-CoV-2, as it is expressed throughout the respiratory tract which may facilitate SARS-CoV-2 entry in the airway and may even lead to cytokine storm when co-expressed with ACE2 [44]. *DPP4* expression was reported to be lower in granulosa cells of women with PCOS in the microarray data analysis [37]. Few other studies reported variable transcript/protein expression of DPP4 in granulosa cells. Braga et al, found no difference in serum DPP4 levels whereas Blauschmidt et al, reported higher plasma DPP4 levels in women with PCOS [45, 46]. Higher DPP4 mRNA levels were observed in KGN granulosa cells after androgen treatment. These are opposing to our results we found lower levels of *DPP4* transcript in both types of granulosa cells. The DHT/insulin induced rat model of PCOS showed lower *DPP4* mRNA and protein levels in ovarian tissue than that of controls which accorded with our results [47, 48].

C-type lectin receptor, CLEC4M allows anchoring of enveloped virus by identifying the glycoproteins structure, which is important, especially contemplating the SARS-CoV-2 pathophysiology. This receptor expression has been found in the ovary and MGCs by single cell RNA sequencing and flow cytometry [13]. In the present study, both microarray analysis and qRT-PCR data showed comparable transcript levels of *CLEC4M* in MGCs and CGCs of control and PCOS women.

The SARS-CoV-2 hijacks transmembrane proteases for priming of its S protein out of which TMPRSS2 is the primary protease [49]. The expression of essential protease for SARS-CoV-2 entry, TMPRSS2, was not different in the ovary between women with PCOS and controls across microarray datasets. However, we could not detect expression of *TMPRSS2* in MGCs and CGCs of controls and PCOS women. In the same line, few other studies reported extremely low or no expression of TMPRSS2 in the ovary [10, 15, 16]. Moreover, any of the cell types in ovarian tissue; namely, granulosa, stromal, endothelial, perivascular or immune cells did not show any evidence of TMPRSS2 expression which was studied using bulk RNA-seq data from Genotype-Tissue Expression (GTEx) Portal [4]. Expression of any alternate TMPRSS protease family member too was not reported in the same study. The available data regarding TMPRSS2 expression in ovary is therefore inconsistent.

We investigated the expression of alternative proteases cathepsins (CTSB/L) and FURIN as they too can prime the S protein of SARS-CoV-2 [4, 26, 50]. CTSL is known to be involved in expansion of cumulus oocyte complex at the time of ovulation [51]. Microarray studies on granulosa cells obtained from women with PCOS revealed variable data on *CTSL* expression (GSE106724), [37]. We found down-regulated *CTSL* expression in MGCs of women with PCOS. However, oksjoki et al observed comparable levels in the ovarian tissue of women with PCOS as compared to controls [52]. *CTSB* expression has been studied in oocytes and granulosa cells and has been found to be higher in women with PCOS than that of controls [37, 38]. However, we found a significantly lower expression *of CTSB* in MGCs and CGCs from PCOS women. The mice treated with insulin to mimic the ovarian dysfunction imparted by hyperinsulinemia in PCOS have showed lower CTSB transcript and protein levels in ovarian tissue in comparison to controls, similar to our gene expression data [53]. *FURIN* is reported to be expressed in granulosa cells and may be playing role in apoptosis and proliferation of these cells [54, 55]. Microarray data showed that *FURIN* was down-regulated in oocyte of women with PCOS whereas in another study, it was reported to be up-regulated in granulosa cells of women with PCOS [35, 38]. We found comparable expression of *FURIN* in granulosa cells of control and PCOS women. S protein has multiple sites for cleavage by FURIN and it is hypothesized that because of this, SARS-CoV-2 may possess higher membrane fusion capacity than other coronaviruses which encourages further purview to inspect this molecule [56].

Our data shows down-regulation of few SCARFs in MGCs and CGCs from women with PCOS as compared to controls. The current data on association between PCOS and COVID-19 is limited. Though, there are some studies with the notion that women with PCOS may be at higher risk of COVID-19 due to excess androgen, lower vitamin D, other co-morbidities and interplay between these elements and cytokine levels in women with PCOS [27, 57, 58]. However, these studies feature systemic SARS-CoV-2 infection and suggest that the higher susceptibility of women with PCOS to COVID-19 may even depend on specific phenotypes. The extent of infection to different cell types by SARS-CoV-2 can vary in different conditions and we have partly answered the same by dissecting SCARF expression in MGCs and CGCs in ovary of women with PCOS. Also, our data hints that ovarian follicle and oocyte may be at lower risk of infection by SARS-CoV-2, Nonetheless these findings need to be established with higher number of samples. Moreover, the expression of SCARFs in granulosa cells may vary in different phases of menstrual cycle as some of them are regulated by sex hormones [30, 59, 60]. Despite these limiting factors, we believe that delineating SCARFs expression in various cell types and physiological conditions holds prime importance as they are putative candidates for developing diagnostic, therapeutic interventions and to better understand the pathogenesis and prognosis of COVID-19. There are past evidences showing that viruses like Epstein-Barr virus, Hepatitis virus can infect ovary and replicate in ovum hence leading to vertical transmission and may be responsible for either infertility, oocyte apoptosis, ovarian failure, or may even cause chronic inflammation and cause ovarian cancer too [61–63]. Considering these repercussions, it becomes even more important to follow the similar approach like ours, i.e. to unravel the likelihood of infection by SARS-CoV-2 to different cells, tissues and organs.

Our data suggests that risk of ovary getting infected by SARS-CoV-2 is less however, we can’t rule out the possibility that it can still get infected as ovary does express SCARFs. Therefore, further studies are warranted in animals and in infected women to understand effect of SARS-CoV-2 infection to the ovary, oocyte and its plausible influence on fertilization. This information may prove to be crucial to prevent the possible spread of transmission of infection in IVF procedures as oocyte/embryo may be exposed to infection at various steps. The biological transmission of virus at IVF settings is possible from potential sources (other than plausible maternal transmission) such as semen from infected males, liquid nitrogen spills, infected healthcare workers. Appropriate SOPs can be laid down in ART procedure to reduce the risk of oocyte and embryo with SARS-CoV-2 infection. Real time, evidence based studies are required to update these guidelines from time to time to avoid this legitimate concern. Some of the operational changes to be implemented at different levels in the ART procedure which have been described in a sophisticated way by ceschin et al and Choucair et al [22, 41].

Emerging data from ongoing animal model studies may provide more insights about the effects of SARS-Co-2 infection on ovary and events preceding and following fertilization. SCARFs knockout/knockdown animals (with varying combinations of genes) and using organoids can be some intriguing approaches to further delve into roles of SCARFs, susceptibility of different organs to SARS-CoV-2 infection and potential routes of infection.

## Supporting information

The expression of genes was investigated using the Takyon SYBR mastermix (Eurogentec, Europe) and appropriate primers

## Data Availability

All the data is included in the manuscript, figures and tables

## Supplementary material

Supplementary data to this article can be found online at https://doi.org/

## Declarations

### Authors approval

All authors have seen and approved the manuscript

### Availability of data and materials

Not applicable

### Code availability

Not applicable

### Authors’ contributions

SM designed the research, edited and approved the final manuscript. AN and KP performed the experiments, analyzed the data, wrote and edited the manuscript. SJ analyzed the data and edited the manuscript. IH provided patient samples.

### Ethical approval

This study was approved by Ethics committee of ICMR-National Institute for Research in Reproductive Health (ICMR-NIRRH).

### Consent to participate

Informed written consent was obtained from all individual participants included in the study

### Consent for publication

Not applicable

## Acknowledgements

The authors thank all study participants Authors extend their gratitude to Dr. Veena Bangera, Ms. Leena Mukadam (P.D. Hinduja National Hospital and Medical Research Centre) who collected clinical samples. We would also like to thank Ms. Ishita Mehta for technical support. We acknowledge necessary support from NIRRH and Indian Council of Medical Research (NIRRH/RA/1084/06-2021), India.

