## Supplementary material for "OVARIAN GRANULOSA CELLS FROM WOMEN WITH PCOS EXPRESS LOW LEVELS OF SARS-COV-2 RECEPTORS AND CO-FACTORS": The expression of genes was investigated using the Takyon SYBR mastermix (Eurogentec, Europe) and appropriate primers

**Supplemental Table 1: Primer sequences and PCR conditions for gene expression**

| Gene symbol | Primers for quantitative real time PCR | | Primer annealing temperature |
|---|---|---|---|
| | Forward primer 5' to 3' | Reverse primer 5' to 3' | |
| ACE2 | TCCATTGGTCTTCTGTCACCCG | AGACCATCCACCTCCACTTCTC | 61°C |
| TMPRSS2 | CCTCTAACTGGTGTGATGGCGT | TGCCAGGACTTCCTCTGAGATG | 61°C |
| | TCGATTCTTGCCAGGGTGAC | ATGTGGATTAGCCGTCTGCC | 61°C |
| BSG | GGCTGTGAAGTCGTCAGAACAC | ACCTGCTCTCGGAGCCGTTCA | 60°C |
| CLEC4M | GAGTAACCGCTTCTCCTGGATG | CGCACAGTCTTCATTCCCGCTA | 61°C |
| DPP4 | AAAGGCACCTGGGAAGTCATCG | CAGCTCACAACTGAGGCATGTC | 59°C |
| CTSL | GAAAGGCTACGTGACTCCTGTG | CCAGATTCTGCTCACTCAGTGAG | 61°C |
| CTSB | GCTTCGATGCACGGGAACAATG | CATTGGTGTGGATGCAGATCCG | 59°C |
| FURIN | GCCACATGACTACTCCGCAGAT | TACGAGGGTGAACTTGGTCAGC | 59°C |

**PCR conditions**

| Stage 1 | Stage 2 | | | Stage 3 | Stage 4 |
|---|---|---|---|---|---|
| 95°C 10 mins | 95°C 10s | Annealing Temperature 15s | 72°C 20s | 72°C 40s | 4°C |
| X 1 | X 40 | | | X 1 | |

*ACE2, angiotensin converting enzyme II; BSG, Basigin; DPP4, Dipeptidyl-peptidase 4; CLEC4M, C-Type Lectin Domain Family 4 Member M; TMPRSS2, Transmembrane protease, serine 2; CTSB, cathepsin B; CTSL, cathepsin L; FURIN, furin.*
